# Early Life Predictors of Child Development at Kindergarten: A Structural Equation Model using a Longitudinal Cohort

**DOI:** 10.1101/2024.11.06.24315323

**Authors:** Sarah E. Turner, Stephanie Goguen, Brenden Dufault, Teresa Mayer, Piushkumar J. Mandhane, Theo J. Moraes, Stuart E. Turvey, Elinor Simons, Padmaja Subbarao, Meghan B. Azad

## Abstract

**Introduction:** Early child development sets the stage for lifelong health. Identifying early life factors related to child development can help guide programs and policies to bolster child health and wellbeing. The objective of this research was to examine how a broad range of predictors, measured prenatally to the third year of life, are related to child development at kindergarten.

**Methods:** We linked survey data from the Manitoba site of the CHILD Cohort Study with data from the Early Development Instrument (EDI) assessment, completed in kindergarten by the Manitoba public school system (n=442 children). The EDI measures five domains of development (ex. language, physical), scored to indicate the bottom 10% (i.e. ‘vulnerable’) of the population on one or more domains. Using structural equation modelling, we grouped 23 predictors of child development into six latent factors including prenatal exposures, child health and lifestyle, family stress, and socioeconomic status. We examined the associations between each latent factor and EDI vulnerability.

**Results:** Overall, 20.1% of children were vulnerable on one or more EDI domains. Higher family stress at 1 year and 3 years was related to a 0.20 (p-value ≤ 0.001) and 0.33 (p-value ≤ 0.001) standardized increase of EDI vulnerability. Higher socioeconomic status was related to a - 0.26 (p-value =0.01) standardized increase of EDI vulnerability, and this link was partially mediated through family stress at three years (10.6% mediated). Prenatal exposures (e.g. maternal diet quality), as well as child health and lifestyle factors (e.g. weekday sleep) were not related to EDI vulnerability.

**Conclusions:** Supporting parental mental health throughout early life, universal screening for early life stress, as well as targeting programs and supports for those living with low SES appear to be priority areas that could help to improve early child development.

## Introduction

Early child development is critical for later life mental and physical health, learning and behaviour (Black et al., 2021; Brinkman et al., 2013; Guhn et al., 2016). Investing in early childhood development yields the largest economic returns compared to any other time period or life stage (Heckman, 2006); it prevents later life challenges and also promotes health equity by providing a standard foundation of health to all children. The theory of the Developmental Origins of Health and Disease, posits that early life exposures and experiences play a critical role in laying the foundation for health and well-being throughout the lifespan (Baird et al., 2017; Barker, 2007). For example, higher child development scores at school entry are related to lower social emotional problems and lower likelihood of being overweight in adolescence (Black et al., 2021). Further to this, self-reported wellbeing in adolescence is related to lower levels of depression, anxiety and relationship problems in adulthood (Kansky et al., 2016).

The Early Development Instrument (EDI) is an internationally-recognized, standardized tool for assessing child development and readiness to learn in kindergarten (Janus et al., 2007). The EDI consists of five domains: 1) physical health and well-being, 2) social competence, 3) emotional maturity, 4) language and thinking skills and 5) communication skills and general knowledge. Each domain has established cut offs to indicate if the child is vulnerable or not (i.e. in the bottom 10^th^ percentile of the population). In Canada, individual provinces have assessed early child development using the EDI since 2005. Between 2010 and 2019, approximately 30% of children in Manitoba had vulnerable scores on one or more EDI domains (Healthy Child Manitoba, 2019). Understanding early life experiences that predict EDI vulnerability can help governments and organizations identify target areas for intervention to help support positive childhood development.

Previous research has identified several early life factors related to poorer EDI scores including maternal depression, lower family socioeconomic status, having a teen mother, maternal smoking during pregnancy and poorer child overall health (Brownell et al., 2016; Chittleborough et al., 2016; Janus & Duku, 2007; Wall-Wieler et al., 2020). However, these studies are limited by studying single predictors, examining only one time point, grouping all predictors into one model or using administrative data which only captures information from service use records. To address these knowledge gaps, we harnessed the rich survey data from the CHILD Cohort and included information on 23 predictors including prenatal exposures, child health and lifestyle, family stress, and socioeconomic status. We grouped predictors into categories, instead of examining them individually, to help gain a more holistic picture of the child’s exposures (Browne et al., 2015). Our objective was to determine how different categories of exposures during the prenatal period and throughout the first three years of life are related to child development at kindergarten, as measured by the EDI.

## Methods

### Study Population

We used a subset of data from the Canadian Healthy Infant Longitudinal Development (CHILD) Cohort Study, a national population-based birth cohort beginning in 2008 and recruiting from four centers across Canada; Toronto, Manitoba (including participants from Winnipeg, Morden and Winker), Edmonton and Vancouver. Details of the cohort can be found elsewhere (Subbarao et al., 2015). The current analysis was limited to the Manitoba site (n=998). CHILD data were linked with data from the Government of Manitoba (GOM) using Personal Health Identification Numbers. Every other year, the GOM routinely collects Early Development Instrument (EDI) data during kindergarten. The biennial data collection of the EDI limits the sample for the current study to approximately half that of the Manitoba CHILD site. CHILD participants who consented to administrative data linkage and had complete EDI data were included in the study (n = 442) (**Supplementary Figure 1**). Informed written consent was obtained by all participating parents prior to data collection and this study was approved by the Human Research Ethics Boards at McMaster University and University of Manitoba.

### The Early Development Instrument

The EDI is a 104 item questionnaire that measures children’s readiness for school at kindergarten across five domains of child development; 1) physical health and well-being, 2) social competence, 3) emotional maturity, 4) language and thinking skills and 5) communication skills and general knowledge (Janus et al., 2007). Each domain has a score ranging from 0 to 10 with higher scores indicating better development. The EDI categorizes children into one of four categories in each domain based on percentile cutoffs in the population: top (highest 25%); middle (middle 50%), at risk (bottom 25%-10%) and vulnerable (bottom 10%). We used percentile data from the entire population of Canadian children and applied the score cut offs to our CHILD cohort sample (**Supplementary Table 1**). Following this, we derived an outcome variable that classified children as vulnerable or not on one or more EDI domains (i.e. EDI vulnerability) (Janus et al., 2007).

### Latent Factors Comprising Predictors of Child Development

Latent factors are unobserved variables that are calculated and measured by observed variables (Brownell et al., 2016; Rosseel, 2012). In collaboration with early child development experts from the University of Manitoba and the GOM, we developed a theoretical model of predictors of child development at kindergarten using available CHILD data and grouped the individual variables into latent factors based on previous literature and our study team’s expertise. We evaluated a range of potential predictor variables across 8 original latent factors (n=35 variables considered in total) and used model fit statistics to select and refine each latent factor by reclassifying variables into new latent factors or omitting variables entirely when they impeded good fit. This process left us with n=23 total predictor variables classified into 6 latent factors in the final model. The latent factors included: (1) prenatal risk behaviours; (2) family stress at one year; (3) child health at one year; (4) child health and lifestyle at three years; (5) family stress at three years; and (6) socioeconomic status. Descriptions of the latent factors are below.

### Description of Latent Factors

“Prenatal risk behaviours” is comprised of data from prenatal questionnaires. Mother’s smoking status during pregnancy was a binary variable (no smoking compared to any smoking during pregnancy). Maternal stress was measured using the Perceived Stress Scale (PSS), a widely used 10-item instrument for measuring perception of stress in the last month (Cohen, 1994; Cohen et al., 1983). The PSS ranges from 0 to 40, with higher scores indicating more stress. Maternal depression was measured using the Centre for Epidemiologic Studies Depression (CES-D) scale, a 20-item measure which asks caregivers to rate their experiences of symptoms associated with depression over the last week (Radloff, 1977). CES-D scores range from 0 to 60, with higher scores indicating more depressive symptoms. All stress and depression variables were used as continuous measures. Mother’s diet was collected using the updated Healthy Eating Index (HEI) 2010 total score (range 0 to 130). HEI is a measure of diet quality that meets standards of the United States dietary guidelines using 12 components; higher scores indicate better diet quality (Guenther et al., 2014).

“Family stress at one year” is comprised of data from postnatal questionnaires at six months and one year. These included maternal stress (using the PSS) and depression (using the CES-D) at six months, and parenting stress at one year. Parenting stress was measured using the Parent-Child Dysfunctional Interaction (P-CDI) sub-scale from the Parenting Stress Index. The P-CDI scale is a 12-items parent-reported measure of parent satisfaction with the interactions with their child (Abidin, 1990). The scale ranges from 12 to 60; higher scores indicate more parenting stress between the parent and child. All stress and depression variables were used as continuous measures.

“Child health at one year” is comprised of data from hospital birth charts, survey questionnaires at birth and one year, and a clinical assessment at one year. Weight gain was calculated as the change in weight for age z-scores from birth to one year and characterized into a binary variable (weight gain velocity ≤ 0.67, or weight gain velocity > 0.67 (Azad et al., 2018)). Atopic conditions were characterized into a binary variable (no atopic conditions compared to one or more) using clinical and questionnaire data asking about: child wheezing, atopic dermatitis (physician diagnosis) and atopic conditions to food (using a skin prick test). Smoking in the home was defined as anyone smoking in the home at one year of age (binary). Number of emergency room (ER) visits were measured in the first year of life and categorized as none, one, two, or three or more.

“Child health and lifestyle at three years” is comprised of data from the one and a half to three-year questionnaires. Categories of fruit, vegetables, and sugar-sweetened beverages were created based on questions asked in the child food frequency questionnaire at three years. Fruits and vegetables were combined and dichotomized as less than or equal to five servings per day and greater than five servings per day. Sugar sweetened beverages was dichotomized as no servings or any servings per day. Sleep was derived from combining number of hours of night sleep and nap time durations during the weekday from the child three-year questionnaire. Number of ER visits were measured between one and a half and three years of age and categorized as none, one, two, or three or more.

“Family stress at three years” is comprised of data from the three-year questionnaires. These included maternal stress (using the PSS), maternal depression (using the CES-D) and parenting stress (using the P-CDI). All variables were used as continuous measures.

“Socioeconomic status” is comprised of data from the prenatal questionnaires. Total household income was categorized as < $80,000 or ≥ $80,000. Marital status was defined as married/common law or single/never married/divorced/separated. Education was categorized as no post-secondary degree or completed a post-secondary degree. Since most mothers in the CHILD cohort completed some form of university, we were unable to use more granular categories for education. Perceived socioeconomic status was measured using a picture of a ladder with the top of the ladder being defined as people who have the highest standing in their community, and the bottom of the ladder being defined as people who have the lowest standing in their community. Participants scored where they believed they best fit within the community ladder (Operario et al., 2004).

### Statistical analysis

#### Step 1: Exploring Predictors of Child Development

Characteristics of the entire population (n=442) were stratified by EDI vulnerability. We used univariate logistic regression to determine associations between each of the 23 early-life predictors and EDI vulnerability.

#### Step 2: Developing Latent Factors of Predictors of Child Development

Structural equation modelling (SEM) was used to develop each latent factor by combining early-life predictors. We tested the goodness of fit of the latent factors using four model fit statistics: Confirmatory Factor Index (CFI), Tucker Lewis Index (TLI), Root Mean Square Error Approximation (RMSEA) and Standardized Root Mean Square Residual (SRMR). CFI and TLI are goodness of fit statistics with values ranging between 0 and 1; values ≥ 0.90 are considered good fit (Hu & Bentler, 1999). RMESA and SRMR are badness of fit statistics with values ranging between 0 and 1; values of ≤ 0.10 are considered good fit (Hu & Bentler, 1999; Steiger, 2007). To improve model fit for some latent factors, we used modification indices to select additional model parameters including residual covariance or regression between two variables. Models were adjusted for: child sex (male or female, from the GOM dataset), child age at EDI assessment (continuous in months, from the GOM dataset), maternal race (Caucasian or other, from the CHILD dataset) and older siblings (none or one or more, from the CHILD dataset).

#### Step 3: Testing Latent Factors in a Predictive Model

SEM was performed to understand the relationship between each latent factor and EDI vulnerability using the diagonally weighted least squares estimate for categorical and continuous predictors (Li, 2021). In the case of ordinal and dichotomous outcomes, SEM uses a probit regression approach and assumes that categorical variables have an underlying normal distribution, and therefore estimated path coefficients can be interpreted as a regular linear effect. Standardized coefficients from each SEM were reported to facilitate comparison between models and estimate the relative importance of each latent factor in predicting EDI vulnerability. Standardized coefficients range from 0 to 1, with larger coefficients indicating a stronger relationship with the outcome (Grace & Bollen, 2005). To further understand the pathways in which early childhood exposures are related to child development, we ran two mediation models using latent factors that were significantly related to EDI vulnerability. We tested if the latent factors of family stress at one year or family stress at three years mediated (explained) the relationship between socioeconomic status and EDI vulnerability. A similar mediation model was tested in a previous analysis and found family stress to strongly mediate the relationship between socioeconomic status and the language and cognitive development EDI domain, thus supporting our hypothesized model (Brownell et al., 2016). Analyses were performed using RStudio(RStudio Team, 2022) and R(R Core Team, 2022) (R version 4.2.1 (2022-06-23 ucrt)) using the lavaan package.

## Results

In our CHILD Study sample, 8.4% of children were vulnerable on physical health and wellbeing; 5.9% were vulnerable on social competence, 9.0% were vulnerable on emotional maturity, 6.6% were vulnerable on language and thinking skills and 5.4% were vulnerable on communication skills and general knowledge (**Supplementary Table 1**). Overall, 20.1% (89/442) were vulnerable on one or more domains (**Table 1**).

**Table 1:**
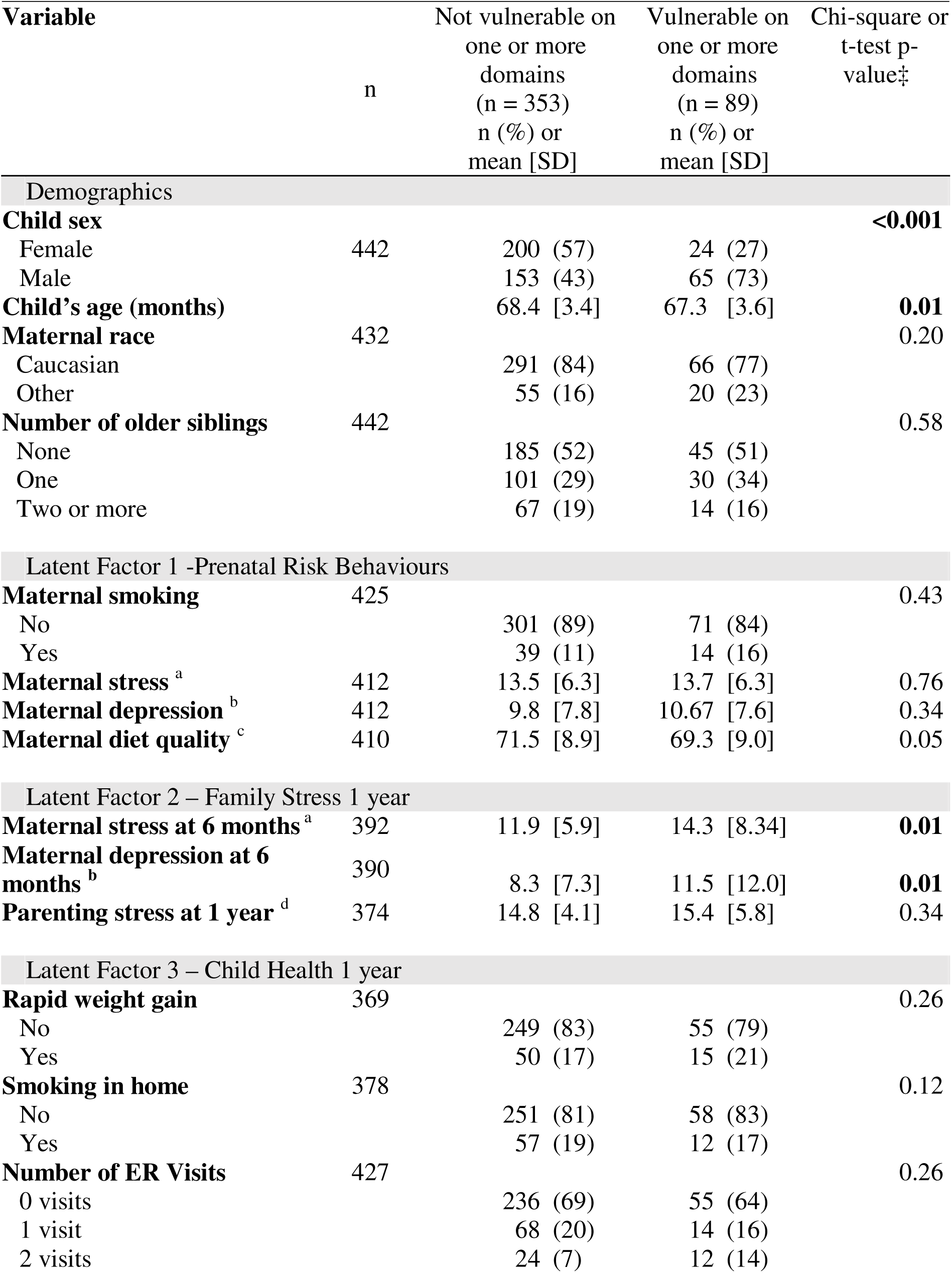
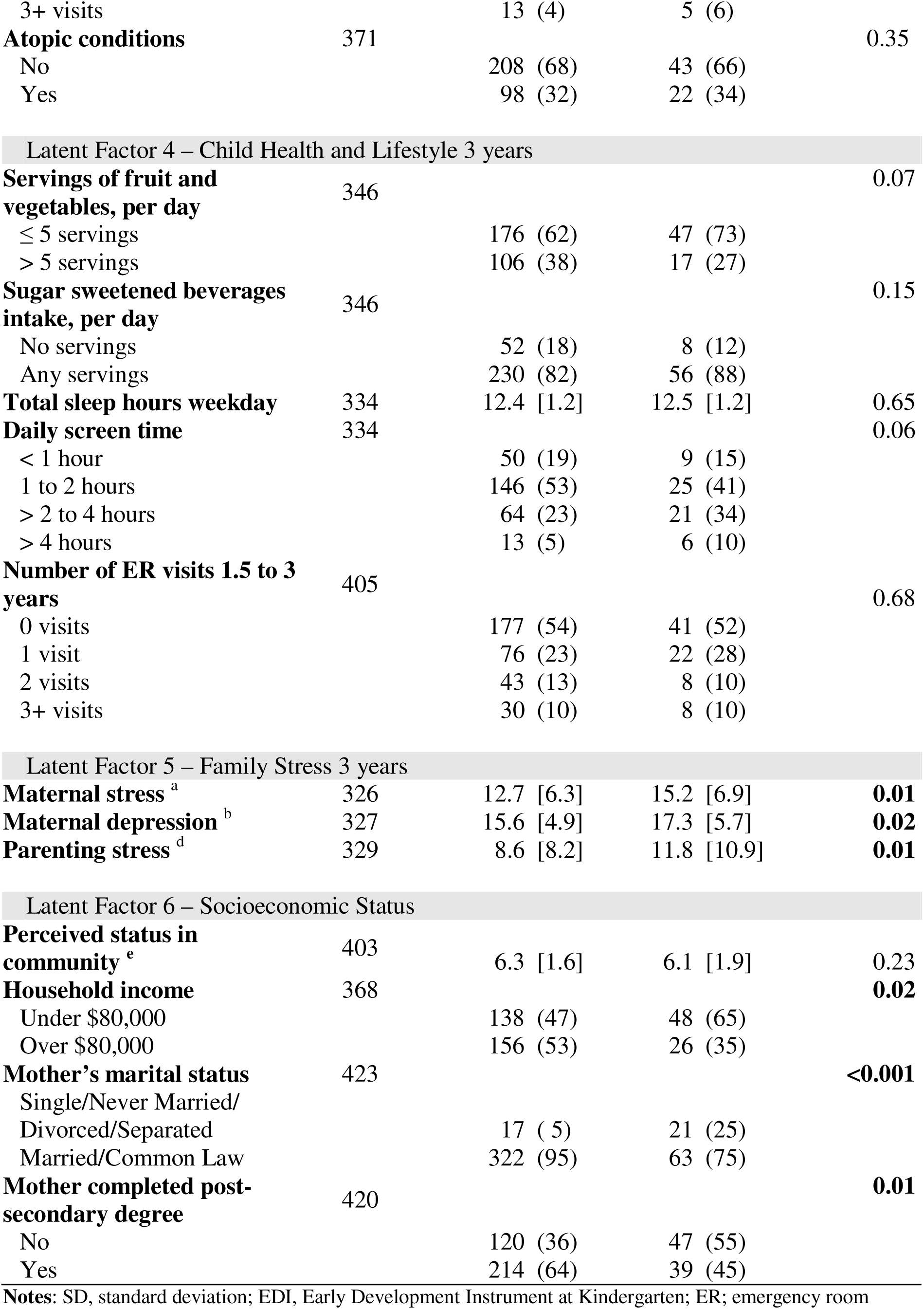
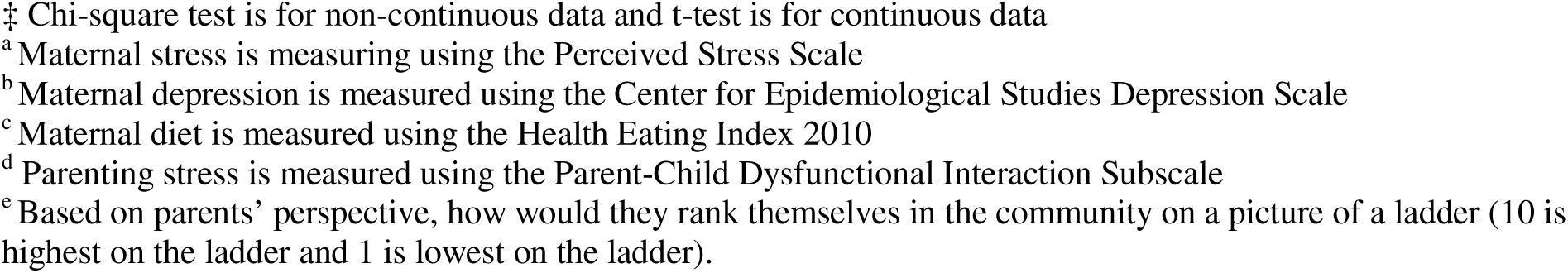
Demographics of Manitoba CHILD Cohort Study Participants, Stratified by EDI Vulnerability at Kindergarten.

| Variable | n | Not vulnerable on one or more domains (n = 353)<br>n (%) or mean [SD] | Vulnerable on one or more domains (n = 89)<br>n (%) or mean [SD] | Chi-square or t-test p-value<sup>‡</sup> |
| --- | --- | --- | --- | --- |
| Demographics |  |  |  |  |
| **Child sex** |  |  |  | **<0.001** |
| Female | 442 | 200 (57) | 24 (27) |  |
| Male |  | 153 (43) | 65 (73) |  |
| **Child's age (months)** |  | 68.4 [3.4] | 67.3 [3.6] | **0.01** |
| **Maternal race** | 432 |  |  | 0.20 |
| Caucasian |  | 291 (84) | 66 (77) |  |
| Other |  | 55 (16) | 20 (23) |  |
| **Number of older siblings** | 442 |  |  | 0.58 |
| None |  | 185 (52) | 45 (51) |  |
| One |  | 101 (29) | 30 (34) |  |
| Two or more |  | 67 (19) | 14 (16) |  |
| Latent Factor 1 -Prenatal Risk Behaviours |  |  |  |  |
| **Maternal smoking** | 425 |  |  | 0.43 |
| No |  | 301 (89) | 71 (84) |  |
| Yes |  | 39 (11) | 14 (16) |  |
| **Maternal stress<sup>a</sup>** | 412 | 13.5 [6.3] | 13.7 [6.3] | 0.76 |
| **Maternal depression<sup>b</sup>** | 412 | 9.8 [7.8] | 10.67 [7.6] | 0.34 |
| **Maternal diet quality<sup>c</sup>** | 410 | 71.5 [8.9] | 69.3 [9.0] | 0.05 |
| Latent Factor 2 – Family Stress 1 year |  |  |  |  |
| **Maternal stress at 6 months<sup>a</sup>** | 392 | 11.9 [5.9] | 14.3 [8.34] | **0.01** |
| **Maternal depression at 6 months<sup>b</sup>** | 390 | 8.3 [7.3] | 11.5 [12.0] | **0.01** |
| **Parenting stress at 1 year<sup>d</sup>** | 374 | 14.8 [4.1] | 15.4 [5.8] | 0.34 |
| Latent Factor 3 – Child Health 1 year |  |  |  |  |
| **Rapid weight gain** | 369 |  |  | 0.26 |
| No |  | 249 (83) | 55 (79) |  |
| Yes |  | 50 (17) | 15 (21) |  |
| **Smoking in home** | 378 |  |  | 0.12 |
| No |  | 251 (81) | 58 (83) |  |
| Yes |  | 57 (19) | 12 (17) |  |
| **Number of ER Visits** | 427 |  |  | 0.26 |
| 0 visits |  | 236 (69) | 55 (64) |  |
| 1 visit |  | 68 (20) | 14 (16) |  |
| 2 visits |  | 24 (7) | 12 (14) |  |
| 3+ visits |  | 13 (4) | 5 (6) |  |
| **Atopic conditions** | 371 |  |  | 0.35 |
| No |  | 208 (68) | 43 (66) |  |
| Yes |  | 98 (32) | 22 (34) |  |
| Latent Factor 4 – Child Health and Lifestyle 3 years |  |  |  |  |
| **Servings of fruit and vegetables, per day** | 346 |  |  | 0.07 |
| ≤ 5 servings |  | 176 (62) | 47 (73) |  |
| > 5 servings |  | 106 (38) | 17 (27) |  |
| **Sugar sweetened beverages intake, per day** | 346 |  |  | 0.15 |
| No servings |  | 52 (18) | 8 (12) |  |
| Any servings |  | 230 (82) | 56 (88) |  |
| **Total sleep hours weekday** | 334 | 12.4 [1.2] | 12.5 [1.2] | 0.65 |
| **Daily screen time** | 334 |  |  | 0.06 |
| < 1 hour |  | 50 (19) | 9 (15) |  |
| 1 to 2 hours |  | 146 (53) | 25 (41) |  |
| > 2 to 4 hours |  | 64 (23) | 21 (34) |  |
| > 4 hours |  | 13 (5) | 6 (10) |  |
| **Number of ER visits 1.5 to 3 years** | 405 |  |  | 0.68 |
| 0 visits |  | 177 (54) | 41 (52) |  |
| 1 visit |  | 76 (23) | 22 (28) |  |
| 2 visits |  | 43 (13) | 8 (10) |  |
| 3+ visits |  | 30 (10) | 8 (10) |  |
| Latent Factor 5 – Family Stress 3 years |  |  |  |  |
| **Maternal stress <sup>a</sup>** | 326 | 12.7 [6.3] | 15.2 [6.9] | **0.01** |
| **Maternal depression <sup>b</sup>** | 327 | 15.6 [4.9] | 17.3 [5.7] | **0.02** |
| **Parenting stress <sup>d</sup>** | 329 | 8.6 [8.2] | 11.8 [10.9] | **0.01** |
| Latent Factor 6 – Socioeconomic Status |  |  |  |  |
| **Perceived status in community <sup>e</sup>** | 403 | 6.3 [1.6] | 6.1 [1.9] | 0.23 |
| **Household income** | 368 |  |  | **0.02** |
| Under \$80,000 | | 138 (47) | 48 (65) | |
| Over \$80,000 | | 156 (53) | 26 (35) | |
| **Mother's marital status** | 423 |  |  | **<0.001** |
| Single/Never Married/ |  |  |  |  |
| Divorced/Separated |  | 17 (5) | 21 (25) |  |
| Married/Common Law |  | 322 (95) | 63 (75) |  |
| **Mother completed post-secondary degree** | 420 |  |  | **0.01** |
| No |  | 120 (36) | 47 (55) |  |
| Yes |  | 214 (64) | 39 (45) |  |

**Notes:** SD, standard deviation; EDI, Early Development Instrument at Kindergarten; ER; emergency room
‡ Chi-square test is for non-continuous data and t-test is for continuous data
<sup>a</sup> Maternal stress is measuring using the Perceived Stress Scale
<sup>b</sup> Maternal depression is measured using the Center for Epidemiological Studies Depression Scale
<sup>c</sup> Maternal diet is measured using the Health Eating Index 2010
<sup>d</sup> Parenting stress is measured using the Parent-Child Dysfunctional Interaction Subscale
<sup>e</sup> Based on parents' perspective, how would they rank themselves in the community on a picture of a ladder (10 is highest on the ladder and 1 is lowest on the ladder).

### Early Life Factors Related to Child Development

Males were over 3 times more likely than females to be vulnerable on one or more EDI domains (odds ratio (OR): 3.54, 95% confidence interval (CI): 2.14-6.01, **Figure 1**). Children were slightly younger in the vulnerable group (9% decrease in being vulnerable per each additional month of age; OR 0.91 95% CI: 0.85-0.98). Having two or more visits to an ER in the first year of life, compared to no visits, was associated with a 2.02-fold increased odds (95% CI: 1.06-3.76) of EDI vulnerability. Daily screen time of greater than two hours was associated with a 2.02-fold increased odds (95% CI: 1.14, 3.57) of EDI vulnerability. A one standard deviation increase in maternal stress and depression scores at six months or three years and parenting stress scores at three years were all associated with increased odds of EDI vulnerability (increased odds between 39% and 43% at six months and 33% and 45% at three years). Protective sociodemographic factors, including having a household income over $80,000 (OR 0.48; 95% CI: 0.28-0.81), a mother who was married or common law (OR 0.16; 95% CI: 0.08-0.32), or a mother who had completed a post-secondary degree (OR 0.47; 95% CI: 0.29-0.75), were associated with lower EDI vulnerability. A one standard deviation increase in maternal prenatal diet quality was associated with a 21% decreased odds (OR 0.79; 95% CI: 0.62-1.00) of EDI vulnerability. Other hypothesized predictors of child development were not statistically different between the vulnerable and not vulnerable groups (ex. older siblings, weekday sleep duration).

**Figure 1:**
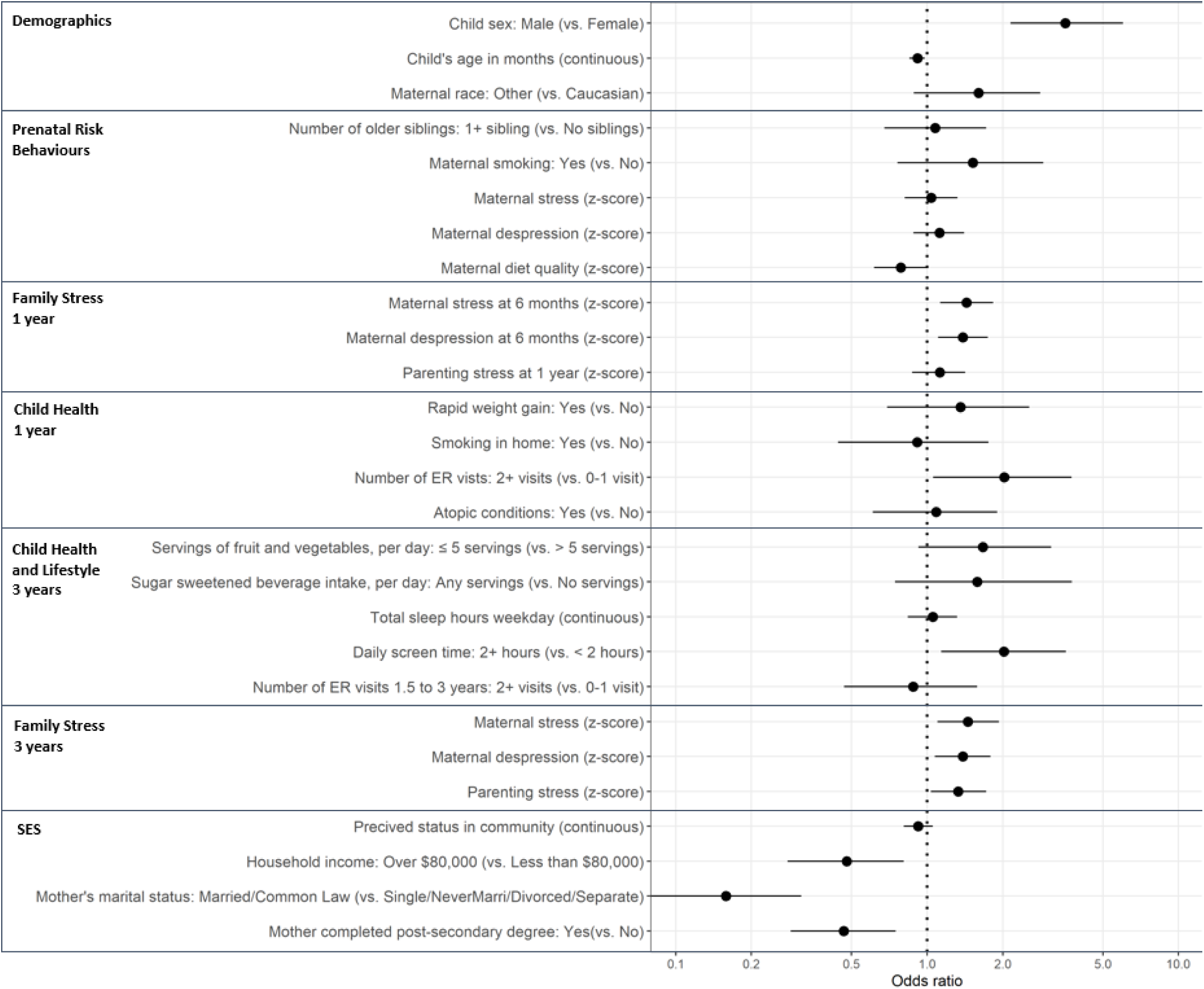
Univariate Logistic Regression Between Early Life Predictors and EDI Vulnerability at Kindergarten in the Manitoba CHILD Cohort Study. **Notes:** Odds ratios predict the odds of being vulnerable on one or more Early Development Instrument (EDI) domains for a one point/category change in the exposure variable. SES, socioeconomic status; ER, emergency room Maternal stress is measuring using the Perceived Stress Scale; Maternal depression is measured using the Center for Epidemiological Studies Depression Scale; Maternal diet is measured using the Health Eating Index 2010; Parenting stress is measured using the Parent-Child Dysfunctional Interaction Subscale; Perceived status in community is based on parents’ perspective, how would they rank themselves in the community on a picture of a ladder (10 is highest on the ladder and 1 is lowest on the ladder). For this table only, maternal depression, maternal stress and parenting stress at one and three years and prenatal healthy eating index are z-score transformed with a mean of zero and a standard deviation of one to allow for more direct comparisons between variables.

### Development of Six Latent Factors to Predict Child Development

All six latent factors passed the thresholds for having good model fit (i.e. CFI or TLI scores ≥ 0.90 and RMSEA and SRMR scores ≤ 0.10; **Table 2**). Most variables (17/23) had standardized loadings that were ≥ 0.32, which fall under Tabachnick and Fidell’s rule of thumb for minimum loading onto a factor (Tabachnick & Fidell, 2013). Some variables were <0.32, but were still included in the model to leverage the richness of the CHILD study data and adhere to our theoretical model of the variables. In addition, variables with low standardized loadings were distributed across the latent factors, rather than grouped onto a single factor, therefore they were not viewed as problematic (Cleare et al., 2018).

**Table 2:**
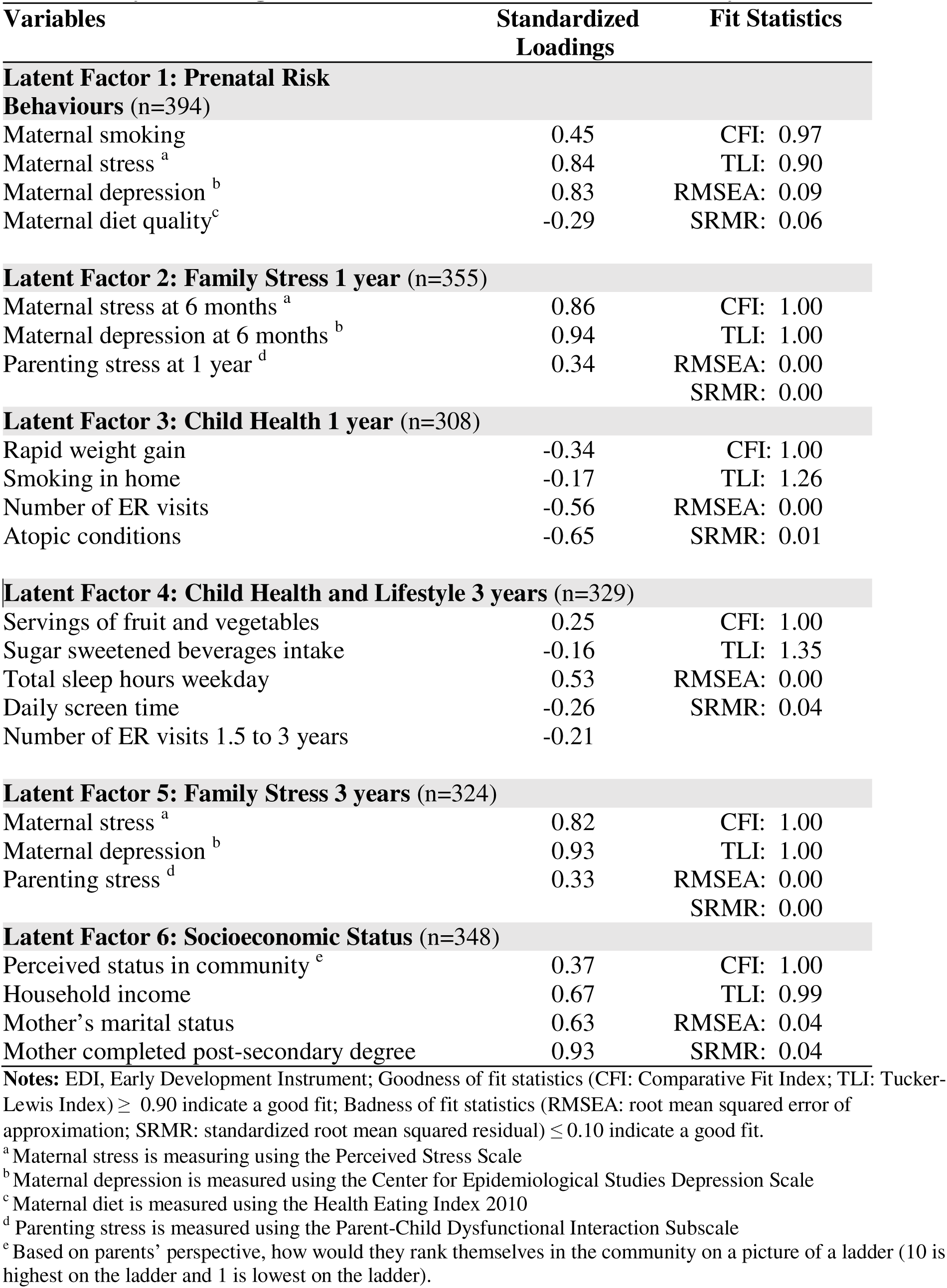
Loadings and Model Fit Statistics for Latent Factors of Predictors of EDI Vulnerability at Kindergarten in the Manitoba CHILD Cohort Study.

| Variables | Standardized Loadings | Fit Statistics |
| --- | --- | --- |
| **Latent Factor 1: Prenatal Risk Behaviours (n=394)** |  |  |
| Maternal smoking | 0.45 | CFI: 0.97 |
| Maternal stress <sup>a</sup> | 0.84 | TLI: 0.90 |
| Maternal depression <sup>b</sup> | 0.83 | RMSEA: 0.09 |
| Maternal diet quality<sup>c</sup> | -0.29 | SRMR: 0.06 |
| **Latent Factor 2: Family Stress 1 year (n=355)** |  |  |
| Maternal stress at 6 months <sup>a</sup> | 0.86 | CFI: 1.00 |
| Maternal depression at 6 months <sup>b</sup> | 0.94 | TLI: 1.00 |
| Parenting stress at 1 year <sup>d</sup> | 0.34 | RMSEA: 0.00 |
|  |  | SRMR: 0.00 |
| **Latent Factor 3: Child Health 1 year (n=308)** |  |  |
| Rapid weight gain | -0.34 | CFI: 1.00 |
| Smoking in home | -0.17 | TLI: 1.26 |
| Number of ER visits | -0.56 | RMSEA: 0.00 |
| Atopic conditions | -0.65 | SRMR: 0.01 |
| **Latent Factor 4: Child Health and Lifestyle 3 years (n=329)** |  |  |
| Servings of fruit and vegetables | 0.25 | CFI: 1.00 |
| Sugar sweetened beverages intake | -0.16 | TLI: 1.35 |
| Total sleep hours weekday | 0.53 | RMSEA: 0.00 |
| Daily screen time | -0.26 | SRMR: 0.04 |
| Number of ER visits 1.5 to 3 years | -0.21 |  |
| **Latent Factor 5: Family Stress 3 years (n=324)** |  |  |
| Maternal stress <sup>a</sup> | 0.82 | CFI: 1.00 |
| Maternal depression <sup>b</sup> | 0.93 | TLI: 1.00 |
| Parenting stress <sup>d</sup> | 0.33 | RMSEA: 0.00 |
|  |  | SRMR: 0.00 |
| **Latent Factor 6: Socioeconomic Status (n=348)** |  |  |
| Perceived status in community <sup>e</sup> | 0.37 | CFI: 1.00 |
| Household income | 0.67 | TLI: 0.99 |
| Mother's marital status | 0.63 | RMSEA: 0.04 |
| Mother completed post-secondary degree | 0.93 | SRMR: 0.04 |

**Notes:** EDI, Early Development Instrument; Goodness of fit statistics (CFI: Comparative Fit Index; TLI: Tucker-Lewis Index) $\geq 0.90$ indicate a good fit; Badness of fit statistics (RMSEA: root mean squared error of approximation; SRMR: standardized root mean squared residual) $\leq 0.10$ indicate a good fit.
<sup>a</sup> Maternal stress is measuring using the Perceived Stress Scale
<sup>b</sup> Maternal depression is measured using the Center for Epidemiological Studies Depression Scale
<sup>c</sup> Maternal diet is measured using the Health Eating Index 2010
<sup>d</sup> Parenting stress is measured using the Parent-Child Dysfunctional Interaction Subscale
<sup>e</sup> Based on parents' perspective, how would they rank themselves in the community on a picture of a ladder (10 is highest on the ladder and 1 is lowest on the ladder).

### Family Stress and Socioeconomic Status Predict Child Development

Adjusted, standardized coefficients modeling the relationships between each of the latent factors and EDI vulnerability are represented in **Figure 2** and **Supplementary Table 2**. All models maintained good model fit as indicated by the fit indices (**Supplementary Table 2**). The prenatal risk behaviours latent factor (comprised of prenatal maternal stress, depressing, smoking and diet), was not significantly associated with being EDI vulnerability at kindergarten (standardized estimate= 0.09, p=0.39). Higher family stress at one year and three years was significantly related to EDI vulnerability: a one standard deviation (SD) increase in family stress at one year was associated with a 0.20 standardized increase in EDI vulnerability (p≤0.001); and the association was even stronger for family stress at three years (standardized estimate = 0.33, p≤0.001). Higher socioeconomic status (SES, latent class comprised of maternal education and marital status, household income and perceived status) was related to lower EDI vulnerability (standardized estimate= −0.26, p= 0.01). Our latent factor measures of child health at one year (standardized estimate= −0.04, p=0.77) and child health and lifestyle at three years (standardized estimate= −0.21, p=0.18) were not statistically related to lower risk of EDI vulnerability at kindergarten.

**Figure 2:**
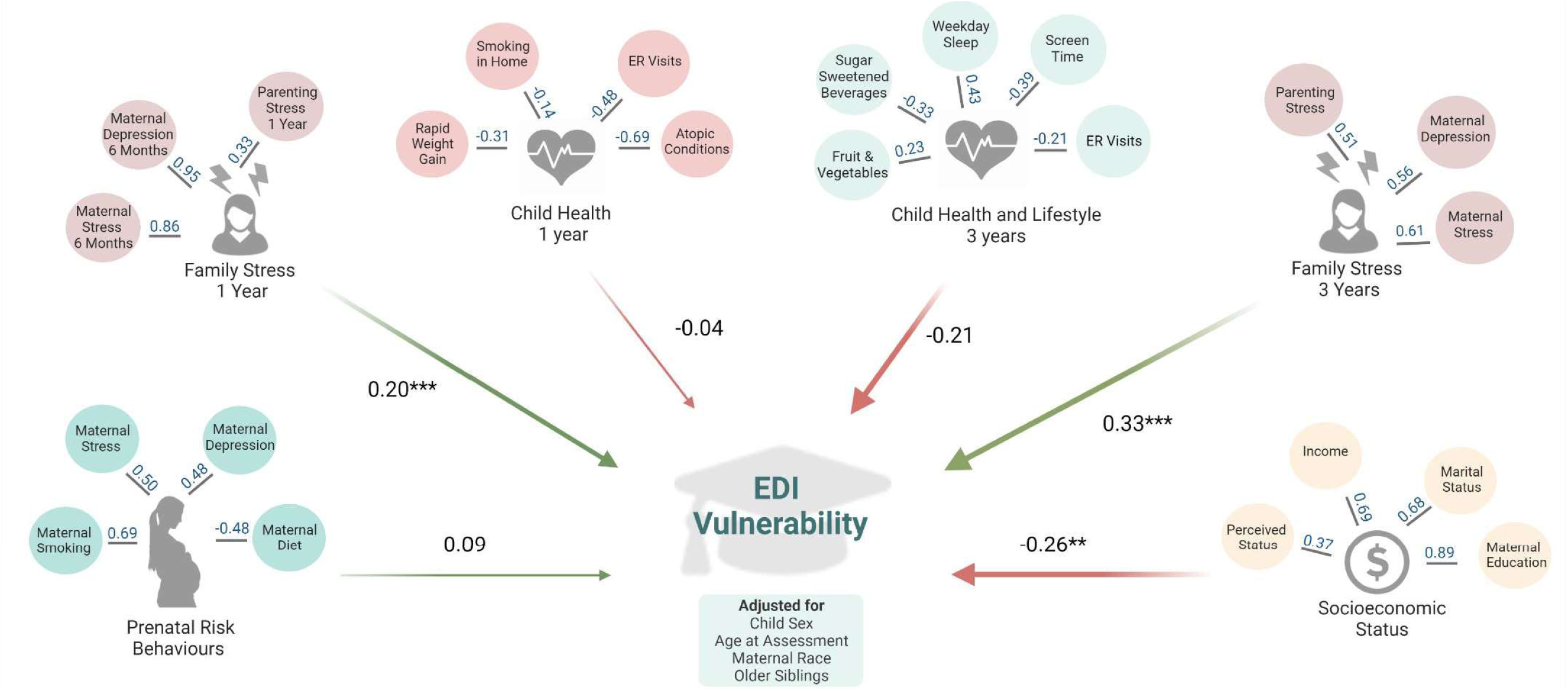
Adjusted Models of Associations between Latent Factors of Early Life Predictors and EDI Vulnerability at Kindergarten in the Manitoba CHILD Cohort Study. **Notes:** All models are adjusted for: child sex, age at Early Development Instrument (EDI) assessment, maternal race, and older siblings. ER, emergency room. This figure represents the results from six separate structural equation models, one for each latent factor. Values in blue indicate factor loadings for each variable onto the latent factor in adjusted regression models. Values in black indicate standardized regression estimates between the latent factor EDI vulnerability. Fit statistics for each structural equation model are in **Supplementary Table 2.** *p:S0.05; **p:S0.01, ***p:S0.001. Created with BioRender.com

### Family Stress at Three Years Mediates the Relationship Between Socioeconomic Status and Child Development

To explore whether the observed association between lower SES and increased EDI vulnerability could be explained by experiencing higher family stress, we performed a mediation analysis using parametric SEM models. After adjustment for covariates, family stress at three years was a significant mediator in the relationship between SES and EDI vulnerability, accounting for 10.6% of the total path standardized estimate (indirect path standardized estimate= −0.05, p ≤ 0.05; **Figure 3** and **Supplementary Table 3**). Family stress at one year did not significantly mediate this relationship (indirect path standardized estimate= −0.03; **Figure 3** and **Supplementary Table 3**); however, it did account for 9.7% of the total path standardized estimate.

**Figure 3:**
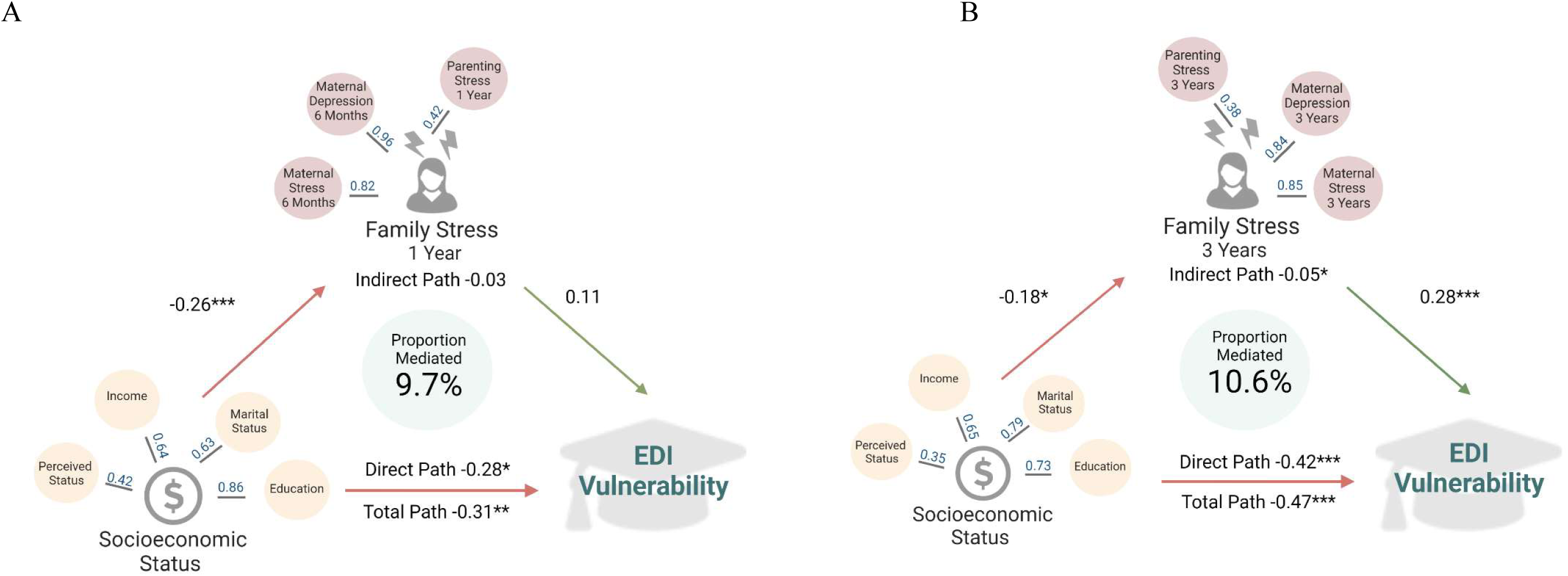
Family Stress at One (A) and Three (B) Years as a Mediator of the Relationship between Socioeconomic Status and EDI Vulnerability at Kindergarten in the Manitoba CHILD Cohort Study. **Notes:** All models are adjusted for: child sex, age at Early Development Instrument (EDI) assessment, maternal race and older siblings. Values in blue indicate factor loadings for each variable onto the latent factor in adjusted regression models. Values in black indicate standardized regression estimates between the exposure and outcome. **Total path** is the change in EDI vulnerability for a one standard deviation increase in the socioeconomic status latent factor. **Indirect path** is the change in EDI vulnerability, through the family stress latent factors. **Direct path** is the change in EDI vulnerability for a one standard deviation increase in socioeconomic status, not through family stress. Fit statistics for each structural equation mediation model are located in **Supplementary Table 3** *p:S0.05; **p:S0.01, ***p:S0.001. Created with BioRender.com

## Discussion

Using structural equation modelling, we evaluated the association of 23 diverse early life factors with child development at kindergarten, measured by the widely-used EDI. Latent factors reflecting family stress at one and three years of life, as well as low SES, were related to EDI vulnerability. Family stress at three years had the largest effect size, and partly explained the link between low SES and EDI vulnerability. Other factors reflecting prenatal risk and early childhood health and lifestyle were relatively unrelated to EDI vulnerability. These results highlight the importance of supporting parents of young children (particularly those of low SES) to minimize family stress throughout the early years to bolster child development.

In this sample of Manitoba children from the CHILD cohort study, 20.1% of participants were classified as vulnerable on one or more EDI domains. Population-level data from Manitoba since 2010 has shown that approximately 30% of children are vulnerable on one or more domains (Healthy Child Manitoba, 2019), indicating that CHILD participants were doing better than the average Manitoba child. This is not surprising given that vulnerable populations are frequently underrepresented in research studies; but it limits the generalizability of our results. Similar to a previous report, being a boy and being younger were associated with EDI vulnerability (Janus & Duku, 2007). Differences in these non-modifiable factors justify conducting sex and age-stratified analysis; however, we did not have adequate sample size in our study.

### Socioeconomic Status and Family Stress are Related to Child Development and Partially Act Along the Same Pathway

Low SES and family stress at one and three years were significant predictors of EDI vulnerability. The detrimental associations of low SES with child development has been established through decades of previous research (Guhn et al., 2020; Letourneau et al., 2011). Single parenthood, maternal and paternal occupation, living in a more disadvantaged area, and lower household income all have significant relationships with EDI vulnerability (Chittleborough et al., 2016; Janus & Duku, 2007). While measures of SES are often not directly modifiable, these results demonstrate the need to provide extra support to children living with lower SES.

The family stress latent factor in our analysis, comprised of maternal stress and depression and parenting stress at one and three years, are modifiable predictors of EDI vulnerability that can be targeted to improve child development. Previous work has established postnatal maternal depression and anxiety as strong predictors of EDI scores (Comaskey et al., 2017; Wall-Wieler et al., 2020). It has been suggested that maternal mental health may interfere with the ability for mothers to be engaged in learning with their child, respond sensitively and consistently, and form secure attachments (Ierardi et al., 2019; Śliwerski et al., 2020; Sohr-Preston & Scaramella, 2006). Infant attachment insecurity has been associated with poorer child executive functioning at kindergarten (Bernier et al., 2015). Furthermore, in the current study, family stress at three years was a significant mediator in the link between SES and early EDI vulnerability at kindergarten. Other studies have examined the interplay between the family environment, SES and child development (Comaskey et al., 2017; Letourneau et al., 2011). In line with our study, previous research shows SES and family stress have both independent and combined effects on child development. Together, this evidence indicates that efforts to reduce poverty, support those living with lower SES and provide services, including universal screening, to reduce family stress and improve family wellbeing, can all be effective in improving child development.

### Prenatal Maternal Health and Postnatal Child Health are Not Related to Child Development

Child health and lifestyle factors at one and three years of age did not emerge as significant predictors of EDI vulnerability. However, the standardized estimate of child health and lifestyle at three years was of similar magnitude to that of family stress at one year (a significant predictor of EDI), indicating that it may still be an important predictor regardless of statistical significance. Contrary to our findings, a previous cross-sectional analysis, found that poorer child health, measured using the Health Utility Index, was significantly related to being vulnerable on one or more EDI domains (Janus & Duku, 2007). The Health Utility Index differs from our measure of health as it is a more comprehensive, standardized assessment and was measured at the time of EDI assessment in the aforementioned cross-sectional analysis.

Prenatal risk behaviours were also not related to EDI vulnerability in the current study. Previous studies have shown that prenatal smoking (Brownell et al., 2016; Chittleborough et al., 2016) and prenatal maternal depression and anxiety (Comaskey et al., 2017) are related to poorer EDI scores. In the current analysis, prenatal maternal depression and stress were not related to EDI vulnerability, however, these same mental health measures at one and three years were related to EDI vulnerability. Previous work has highlighted the potential deleterious effects of exposure to chronic maternal depression and anxiety (Comaskey et al., 2017). While our study did not explicitly measure chronic stress, children exposed to depression or stress at three years may also have been exposed to it at earlier time points, indicating a possible chronic exposure. Further research should focus on the associations of chronic stress during the first three years of life on child EDI scores. Such analysis was beyond the scope of this paper and would require a larger sample size than what was available in the current study.

### Strengths and Limitations

This study is strengthened by combining rich longitudinal survey data from the CHILD cohort with a population-based validated measure of child development. The EDI is teacher reported, removing parental bias on reports of child functioning. Compared to previous research, our study expands the number of predictors evaluated and incorporates groups of predictors into statistically and theoretically coherent latent factors, providing a more holistic approach for examining childhood exposures. The current study is limited by a relatively small sample size compared to previous population-based studies, which may result in type two error whereby significant associations exist, but cannot be detected due to lower power. Additionally, we used complete case analysis and found that missing data were not completely at random, potentially biasing our results (i.e. there was more missing data among those who were vulnerable on one or more EDI domains (**Supplementary Figure 2**)). In addition, the CHILD cohort is comprised of a higher SES profile than the general Manitoba population, limiting the generalizability of our results, especially for the most vulnerable. Finally, while we collected detailed longitudinal data on maternal stress and mental health, we lacked information on their partners’ mental health, which may also impact child development (Kahn et al., 2004).

## Conclusion

This study examined 23 early life predictors of EDI vulnerability to identify areas that would be most beneficial for targeting to improve child development at kindergarten. Living with low SES and experiencing family stress in the first three years of life were significantly associated with EDI vulnerability. Other early life factors, including prenatal risk factors and child health and lifestyle in the first three years of life, did not emerge as significant predictors of child development. This evidence supports the development of programs to promote positive parental mental health and parenting strategies that result in less family stress, and provide resources to those in low socioeconomic environments.

## Data Availability

A list of variables available in the CHILD Cohort Study is available at https://childstudy.ca/for-researchers/study-data/. Researchers interested in collaborating on a project and accessing CHILD Cohort Study data should contact the Study's National Coordinating Centre (NCC) to discuss their needs before initiating a formal request. To contact the NCC, please. More information about data access for the CHILD Cohort Study can be found at https://childstudy.ca/for-researchers/data-access/

