## Supplementary Tables and Figures for "Early Life Predictors of Child Development at Kindergarten: A Structural Equation Model using a Longitudinal Cohort"

**Supplementary Table 1: Proportion of Manitoba CHILD Participants Classified as Vulnerable on the EDI at Kindergarten Based on Canadian Population Cut Offs**

| **EDI Domain** | **Scale  Range** | **Score Cut Off at the Lowest 10% to define “Vulnerable” for Canadian Children** | **Percent of CHILD Participants Categorized as Vulnerable Based on Canadian Cut Offs** |
| --- | --- | --- | --- |
| Physical Health and Well-Being | 0 to 10 | ≤ 7.08 | 8.4% |
| Social Competence | 0 to 10 | ≤ 5.58 | 5.9% |
| Emotional Maturity | 0 to 10 | ≤ 6.00 | 9.0% |
| Language and Thinking | 0 to 10 | ≤ 5.77 | 6.6% |
| Communication Skills and General Knowledge | 0 to 10 | ≤ 4.38 | 5.4% |

Notes: EDI (Early Development Instrument) domain cut offs supplied by the Social Innovation Office of the Government of Manitoba. For all EDI domains, higher scores are better.

**Supplementary Table 2: Adjusted Standardized Regression Estimates and Fit Statistics for Latent Factors of Early Life Predictors and EDI Vulnerability at Kindergarten in the Manitoba CHILD Cohort Study**

| Latent Factors | **Standardized Regression Estimate**  **(Standard Error)** | **p-value** | **Fit Statistics** | |
| --- | --- | --- | --- | --- |
| Latent Factor 1: Prenatal Risk Behaviours (n=393) |  |  |  | |
|  | 0.09 (0.17) | 0.39 | CFI: | 0.99 |
|  |  |  | TLI: | 0.99 |
|  |  |  | RMSEA: | 0.02 |
|  |  |  | SRMR: | 0.04 |
| Latent Factor 2: Family Stress 1 year (n=352) | | | | |
|  | **0.20 (0.01)** | **≤0.001** | CFI: | 1.00 |
|  |  |  | TLI: | 1.00 |
|  |  |  | RMSEA: | 0.00 |
|  |  |  | SRMR: | 0.01 |
| Latent Factor 3: Child Health 1 year (n=306) | | | | |
|  | -0.04 (0.47) | 0.77 | CFI: | 1.00 |
|  |  |  | TLI: | 1.02 |
|  |  |  | RMSEA: | 0.00 |
|  |  |  | SRMR: | 0.04 |
| Latent Factor 4: Child Health and Lifestyle 3 years (n=328) | | | | |
|  | -0.21 (0.75) | 0.18 | CFI: | 1.00 |
|  |  |  | TLI: | 1.01 |
|  |  |  | RMSEA: | 0.00 |
|  |  |  | SRMR: | 0.05 |
| Latent Factor 5: Family Stress 3 years (n=323) | | | | |
|  | **0.33 (0.02)** | **≤0.001** | CFI: | 0.99 |
|  |  |  | TLI: | 1.00 |
|  |  |  | RMSEA: | 0.02 |
|  |  |  | SRMR: | 0.01 |
| Latent Factor 6: Socioeconomic Status (n=348) | | | | |
|  | **-0.26 (0.16)** | **≤0.01** | CFI: | 1.00 |
|  |  |  | TLI: | 1.00 |
|  |  |  | RMSEA: | 0.00 |
|  |  |  | SRMR: | 0.03 |

Notes: EDI, Early Development Instrument; Goodness of fit statistics (CFI: Comparative Fit Index; TLI: Tucker-Lewis Index) ≥ 0.90 indicate a good fit; Badness of fit statistics (RMSEA: root mean squared error of approximation; SRMR: standardized root mean squared residual) ≤ 0.10 indicate a good fit. Results from this table are shown in graphical format in Main Figure 2.

**Supplementary Table 3: Adjusted Standardized Regression Estimates and Fit Statistics for each Mediation Model Predicting EDI Vulnerability at Kindergarten in the Manitoba CHILD Cohort**

| **Latent Factor** | **Mediator** | **Mediation**  **Effects** | **Standardized Regression Estimate (Standard Error)** | **p-value** | **Fit Statistics** | |
| --- | --- | --- | --- | --- | --- | --- |
| Socioeconomic Status  (n= 291) | Family Stress  1 Year | Total | **-0.31 (0.16)** | **≤0.01** | CFI: | 0.95 |
|  |  | Direct | **-0.28 (0.17)** | **≤0.05** | TLI: | 0.97 |
|  |  | Indirect | -0.03 (0.05) | 0.35 | RMSEA: | 0.04 |
|  |  | % Mediated | 9.7% |  | SRMR: | 0.06 |
| Socioeconomic Status  (n= 264) | Family Stress  3 Years | Total | **-0.47 (0.24)** | **≤0.001** | CFI: | 0.93 |
|  |  | Direct | **-0.42 (0.25)** | **≤0.001** | TLI: | 0.95 |
|  |  | Indirect | **-0.05 (0.04)** | **≤0.05** | RMSEA: | 0.05 |
|  |  | % Mediated | **10.6%** |  | SRMR: | 0.09 |

Notes: EDI, Early Development Instrument; Goodness of fit statistics (CFI: Comparative Fit Index; TLI: Tucker-Lewis Index) ≥ 0.90 indicate a good fit; Badness of fit statistics (RMSEA: root mean squared error of approximation; SRMR: standardized root mean squared residual) ≤ 0.10 indicate a good fit. Results from this table are shown in graphical format in Main Figure 3.

**Supplementary Figure 1: CONSORT Flow Diagram for Manitoba CHILD Cohort Study Participants Included in the Present Analysis**

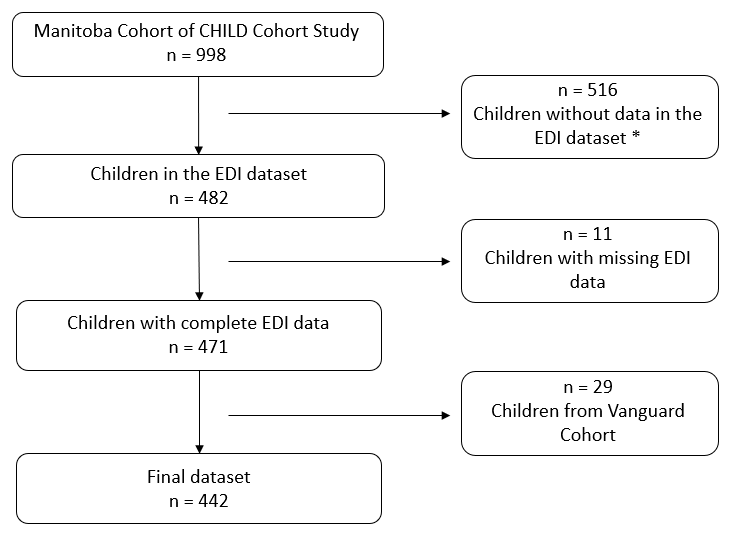

**Note:** The Vanguard cohort was a pilot cohort of the CHILD study that used slightly different questionnaires from the full cohort and therefore was excluded from the analysis. * EDI data are only collected every second year.

**Supplementary Figure 2: Missing Data for Each Variable, Stratified EDI Vulnerability at Kindergarten in the Manitoba CHILD Cohort Study**

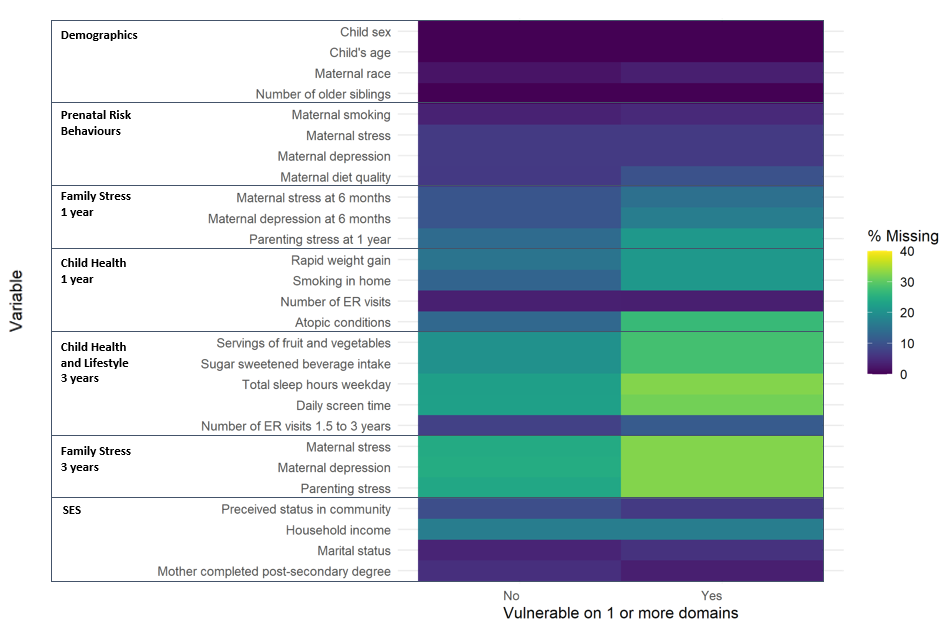

**Note:** EDI, Early Development Instrument; SES, socioeconomic status; ER, emergency room. Maternal stress is measuring using the Perceived Stress Scale; Maternal depression is measured using the Center for Epidemiological Studies Depression Scale; Maternal diet is measured using the Health Eating Index 2010; Parenting stress is measured using the Parent-Child Dysfunctional Interaction Subscale; Perceived status in community is based on parents’ perspective, how would they rank themselves in the community on a picture of a ladder (10 is highest on the ladder and 1 is lowest on the ladder).
